## Supplementary figures for "Spike in asthma healthcare presentations in eastern England during June 2021: a retrospective observational study using syndromic surveillance data"

**Fig S1** Emergency department asthma attendances by age group
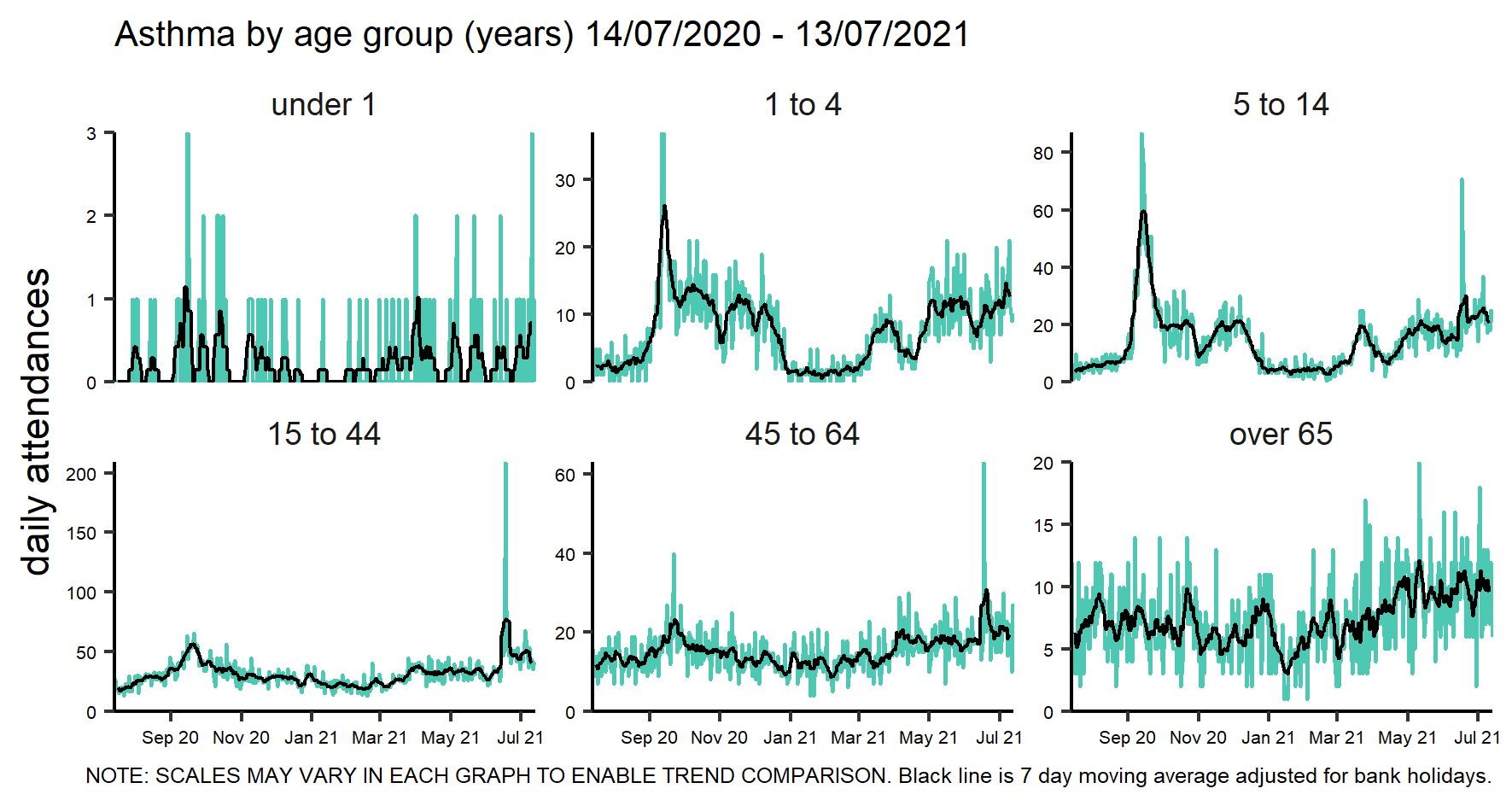

**Fig S2** Emergency department asthma attendances by region of England
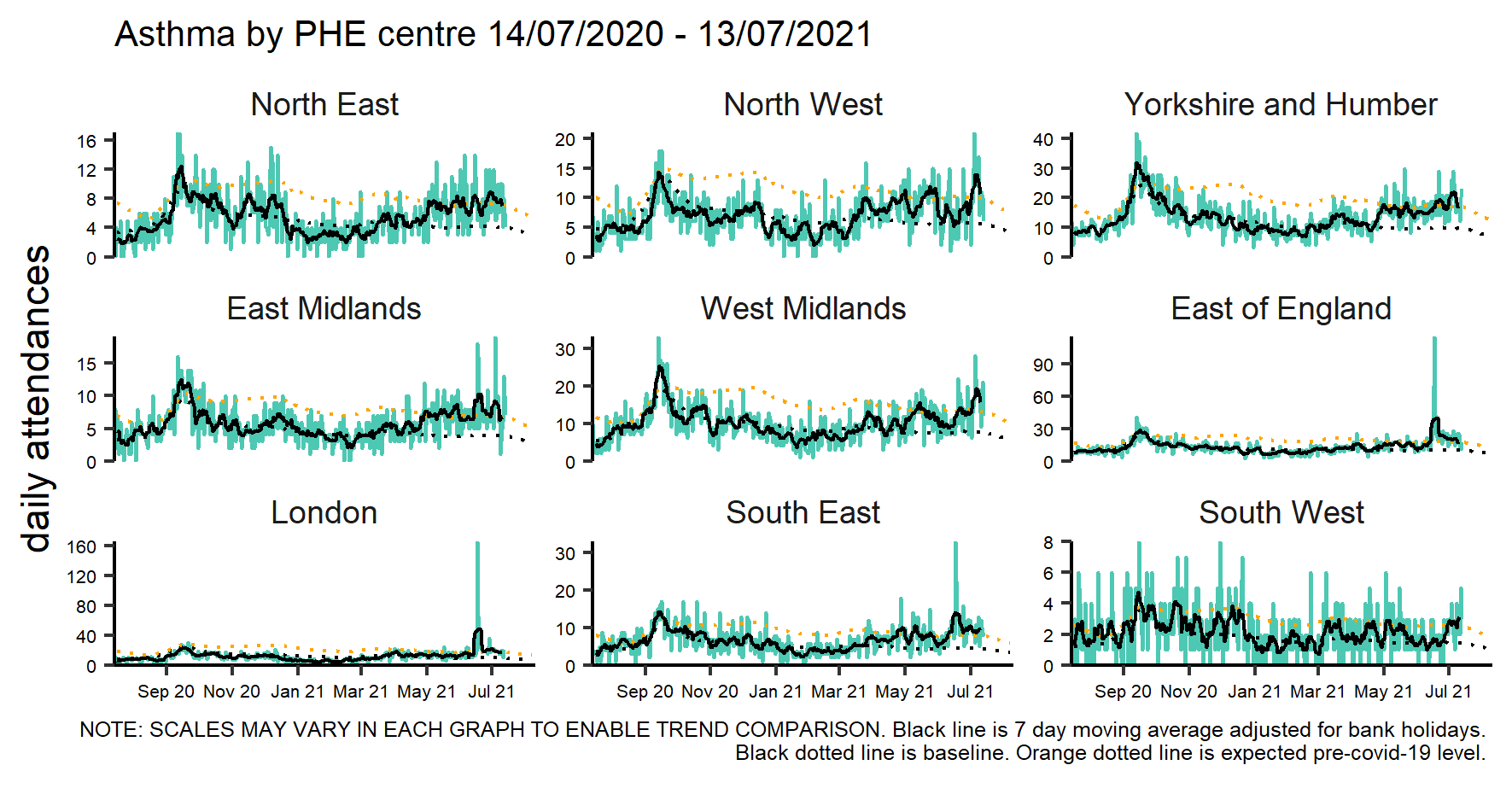

**Fig S3** NHS 111 difficulty breathing calls by age group
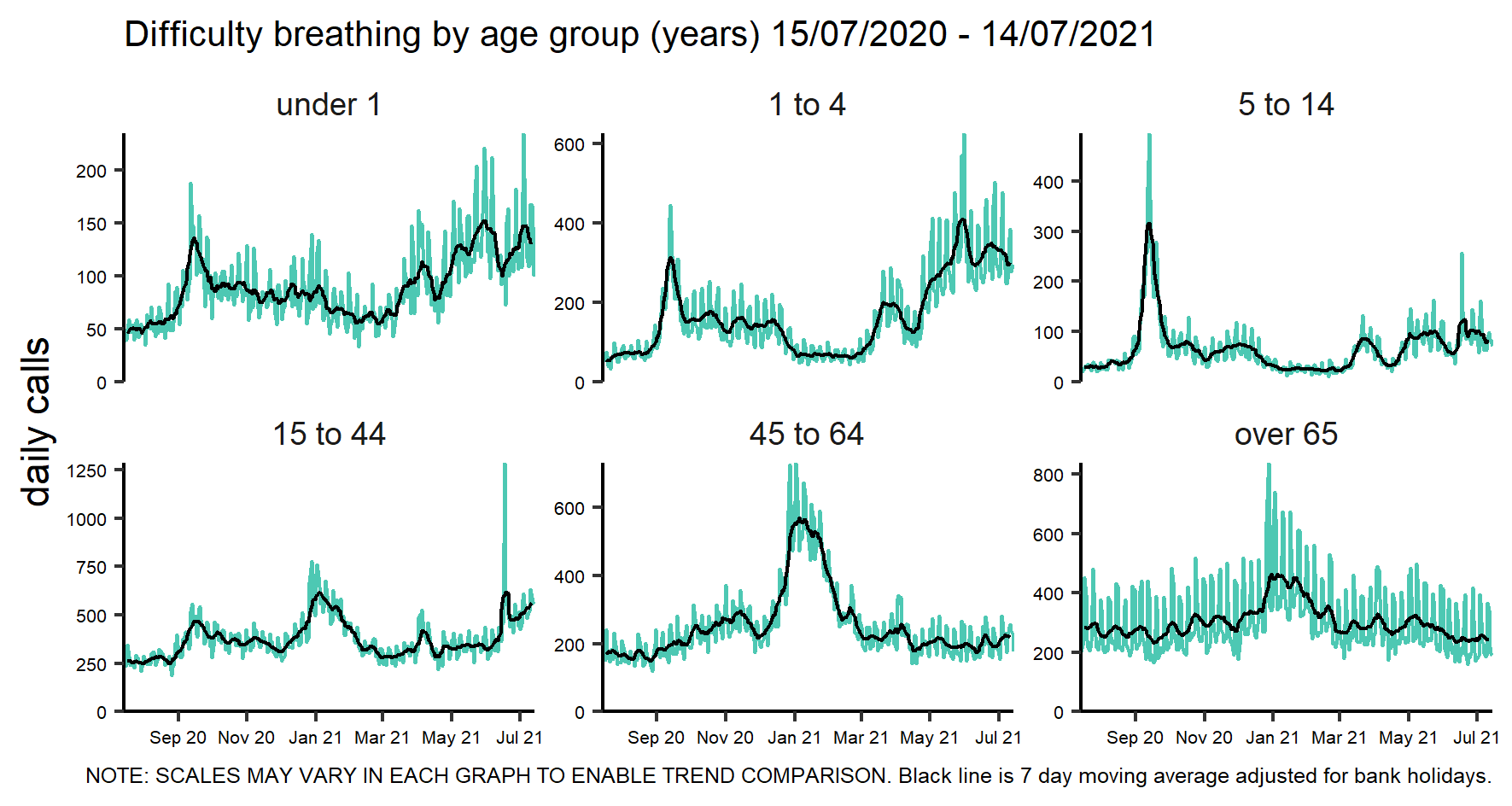

**Fig S4** NHS 111 difficulty breathing calls by region of England
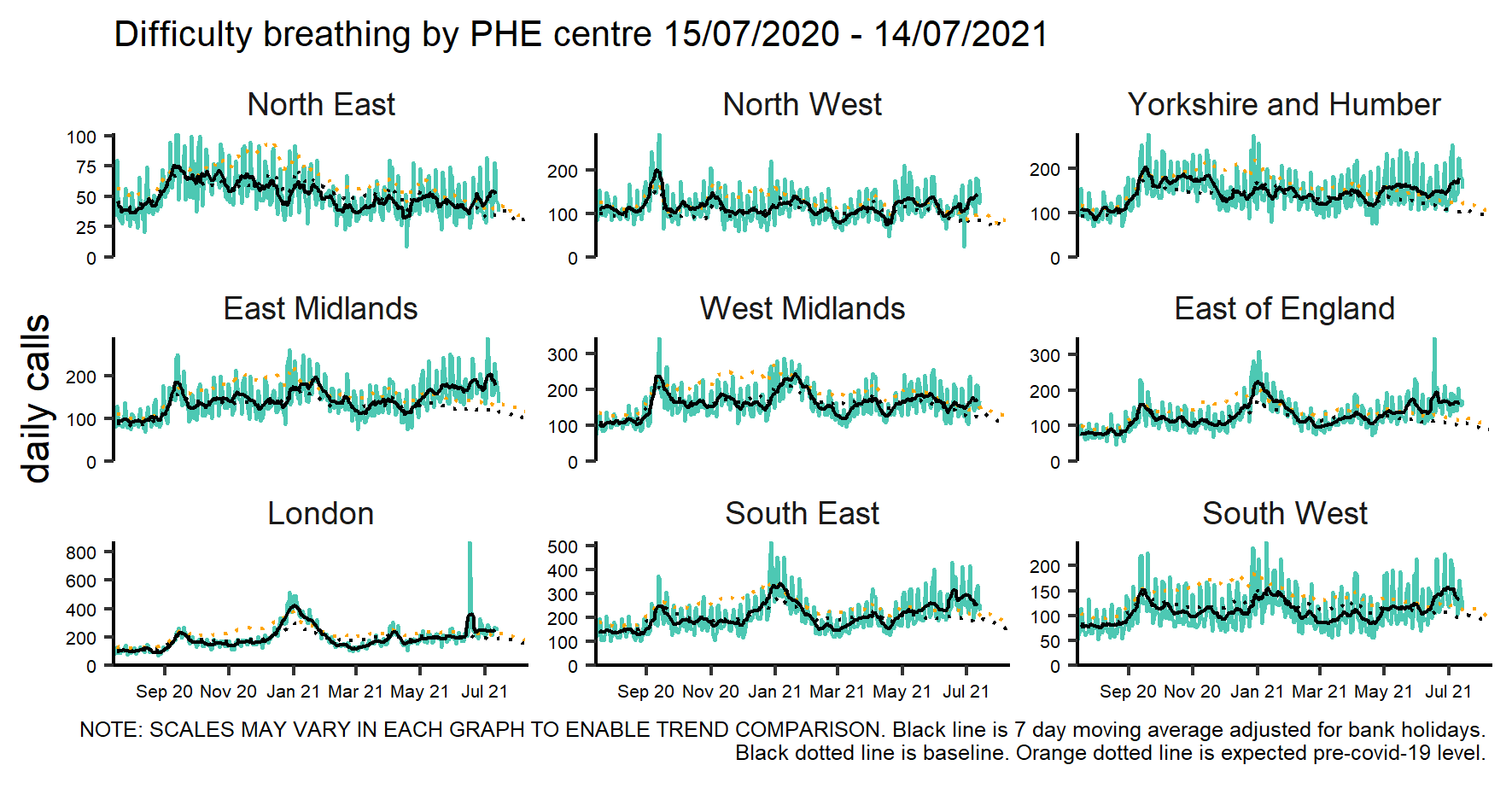

**Fig S5** GP out of hours difficulty breathing/wheeze/asthma contacts by age group
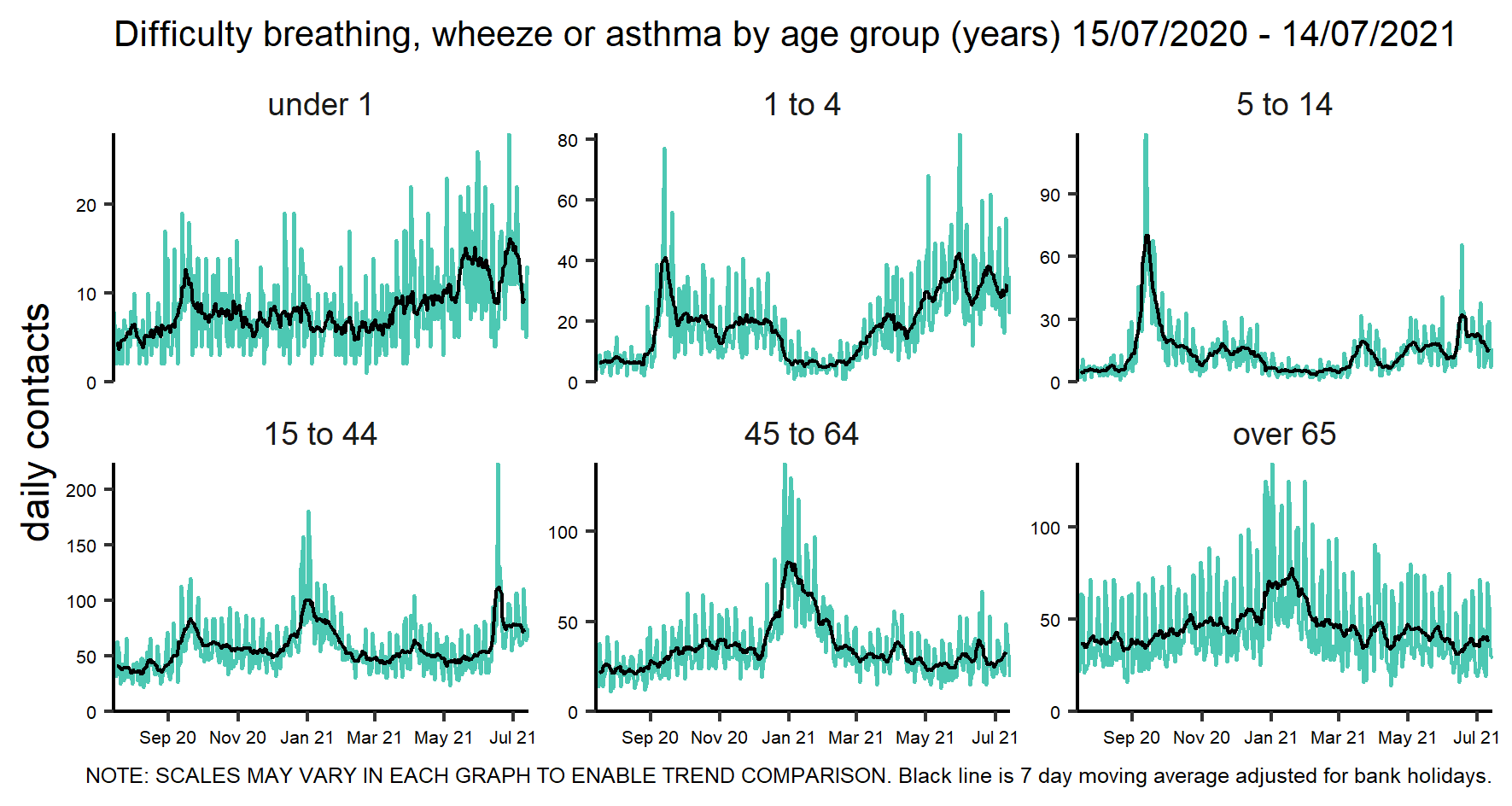

**Fig S6** GP out of hours difficulty breathing/wheeze/asthma contacts by region of England
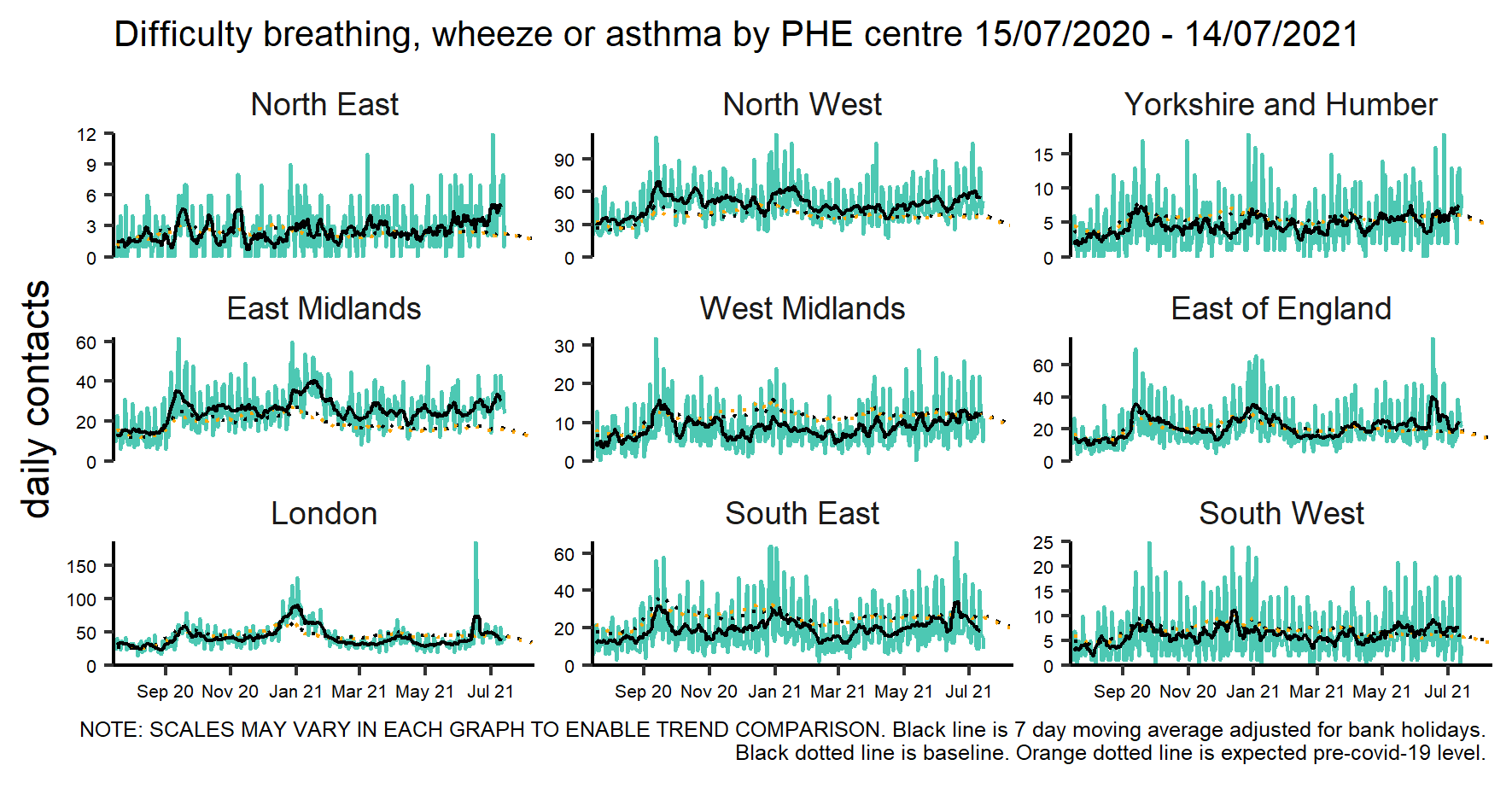

**Fig S7** Ambulance breathing problems calls by region of England
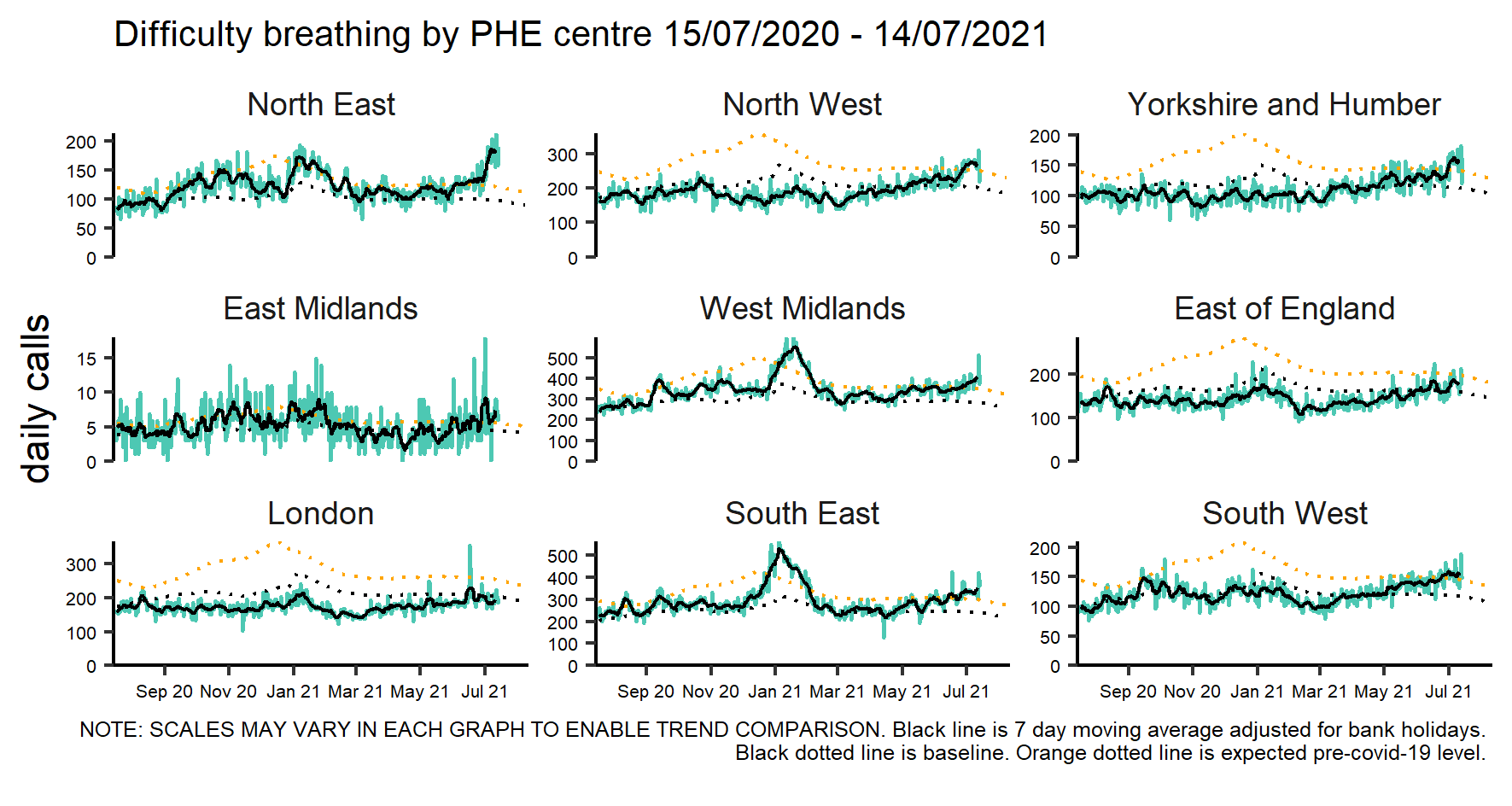
